# Multicentre diagnostic evaluation of OnSite COVID-19 Rapid Test (CTK Biotech) among symptomatic individuals in Brazil and The United Kingdom

**DOI:** 10.1101/2022.09.12.22279847

**Authors:** Caitlin R Thompson, Pablo Muñoz Torres, Konstantina Kontogianni, Rachel L Byrne, LSTM Diagnostic group, Saidy Vásconez Noguera, Alessandra Luna-Muschi, Ana Paula Marchi, Pâmela S Andrade, Antonio dos Santos Barboza, Marli Nishikawara, CONDOR steering group, Richard Body, Margaretha de Vos, Camille Escadafal, Emily Adams, Silvia Figueiredo Costa, Ana I Cubas Atienzar

**Affiliations:** Liverpool School of Tropical Medicine, Centre for Drugs and Diagnostics, Liverpool, UK; LIM-49, Instituto de Medicina Tropical, Faculdade de Medicina da Universidade de São Paulo, São Paulo, Brazil; Departamento de Moléstias Infecciosas e Parasitárias, Faculdade de Medicina da Universidade de São Paulo, São Paulo, Brazil; Department of Epidemiology, School of Public Health of University of São Paulo, São Paulo, Brazil; Centro de atendimento ao colaborador, Hospital das Clínicas da Faculdade de Medicina da Universidade de São Paulo, São Paulo, Brazil; Divisão de Laboratório Central, Hospital das Clinicas, Faculdade de Medicina da Universidade de São Paulo, São Paulo, Brazil; FIND, Geneva, Switzerland; Manchester University NHS Foundation Trust, UK; Global access diagnostics, Bedfordshire, UK

## Abstract

The COVID-19 pandemic has given rise to numerous commercially available antigen rapid diagnostic tests (Ag-RDTs). To generate and share accurate and independent data with the global community, multi-site prospective diagnostic evaluations of Ag-RDTs are required. This report describes the clinical evaluation of OnSite COVID-19 Rapid Test (CTK Biotech, California, USA) in Brazil and The United Kingdom.

A total of 496 paired nasopharyngeal (NP) swabs were collected from symptomatic healthcare workers at Hospital das Clínicas in São Paulo, and 211 NP swabs were collected from symptomatic participants at a COVID-19 drive-through testing site in Liverpool, England. These swabs were analysed by Ag-RDT and results were compared to RT-qPCR.

The clinical sensitivity of the OnSite COVID-19 Rapid test in Brazil was 90.3% [95% Cl 75.1 – 96.7%] and in the United Kingdom was 75.3% [95% Cl 64.6 – 83.6%]. The clinical specificity in Brazil was 99.4% [95% Cl 98.1 – 99.8%] and in the United Kingdom was 95.5% [95% Cl 90.6 – 97.9%]. Analytical evaluation of the Ag-RDT was assessed using direct culture supernatant of SARS-CoV-2 strains from Wild-Type (WT), Alpha, Delta, Gamma and Omicron lineages. Analytical limit of detection was 1.0×10^3^ pfu/mL, 1.0×10^3^ pfu/mL, 1.0×10^2^ pfu/mL, 5.0×10^3^ pfu/mL and 1.0×10^3^ pfu/mL, giving a viral copy equivalent of approximately 2.1×10^5^ copies/mL, 2.1×10^4^ copies/mL, 1.6×10^4^ copies/mL, 3.5×10^6^ copies/mL and 8.7 × 10^4^ for the Ag-RDT, when tested on the WT, Alpha, Delta, Gamma and Omicron lineages, respectively.

This study provides comparative performance of an Ag-RDT across two different settings, geographical areas, and population. Overall, the OnSite Ag-RDT demonstrated a lower clinical sensitivity than claimed by the manufacturer… Sensitivity and specificity from the Brazil study fulfilled the performance criteria determined by the World Health Organisation but the performance obtained from the UK study failed to. Further evaluation of the use of Ag-RDTs should include harmonised protocols between laboratories to facilitate comparison between settings.

## Introduction

To meet the immense diagnostic demand of the COVID-19 pandemic, the use of rapid diagnostic tests for the detection of SARS-CoV-2 antigens (Ag-RDTs) has become a priority. To date, there are currently 321 SARS-CoV-2 Ag-RDTs on the market or in development according to the foundation for new innovative diagnostics (FIND) (date accessed March 2022)[1]. However, clinical evaluation of these Ag-RDTs has been relatively limited and performance results differ greatly between studies[2, 3]. In the UK, the use of Ag-RDTs has been integral to reducing the spread of COVID-19 [4]. However, since April 2022 the UK government has ceased free Ag-RDT testing, now requiring the responsibility of the acquisition, and performance of tests to be placed on the individual.

In Brazil, the national SARS-CoV-2 testing approach has been insufficient in its use of this diagnostic tool in the efforts to contain this pandemic [5]. Many initiatives such as recruiting capacity in university research laboratories and biotechnological enterprises, investments in new laboratory infrastructure and fast-track regulatory measures were launched to scale-up SARS-CoV-2 RT-qPCR testing in Brazil. However, the expansion of the quantitative reverse transcriptase polymerase chain reaction (RT-qPCR) capacity has not been sufficient to control the progress of the pandemic within this country [5].

Despite the commercialisation of several vaccines for SARS-CoV-2, the COVID-19 pandemic is still ongoing due to the vaccine inequity [6], uneven vaccine uptake between populations [7] and the emergence of new SARS-CoV-2 highly transmissible variants [8].

The gold standard for diagnosis of COVID-19 remains the detection of SARS-CoV-2 ribonucleic acid (RNA). However, RT-qPCR requires skilled laboratory scientists, installed capacity and expensive consumables and reagents which can be challenging to implement in low and middle-income countries (LMIC), where the burden of COVID-19 is disproportionately felt. Additionally, turnaround of results of RT-qPCR can take up to one week [9].

In order to continue to meet the challenges of testing capacity, prospective diagnostic evaluation studies across multiple, independent sites are required to determine the accuracy of COVID-19 Ag-RDTs available for purchase to the public.

In this study, OnSite COVID-19 Rapid Test (CTK Biotech) was evaluated against the SARS-CoV-2 diagnostic gold standard RT-qPCR. Testing was undertaken in Brazil and the UK across different settings: on healthcare workers (HCWS) at Hospital das Clínicas, a tertiary-care hospital affiliated with the University of São Paulo (Brazil) and at a National Health Service COVID-19 drive-through community testing centre in Liverpool, UK.

## Methods

### Clinical evaluation

This was a prospective evaluation of consecutive participants enrolled in two different settings:

### Brazil

Healthcare workers (HCW) with suspected COVID-19 symptoms (fever, cough, shortness of breath, tight chest, runny nose, sore throat, anosmia, ageusia, headache and diarrhoea) were enrolled at the HCW service of Hospital das Clínicas in São Paulo from July to October 2021. Ethical approval was obtained from the Hospitalś Ethics Committee with the CAAE number 35246720.0.0000.0068. Informed consent was obtained from all study participants for respiratory samples and clinical data collection.

Participants were clinically evaluated and RT-qPCR for SARS-CoV-2 was performed from combined nasopharyngeal and oropharyngeal swabs (Goodwood Medical Care LTD/(DG) China) following the national standard of care. Following the RT-qPCR swabs, nasopharyngeal (NP) swabs were collected for Ag-RDT testing. The OnSite Ag-RDT was performed at the point-of-care by HCW following manufacturer’s instructions for use (IFU).

For SARS-CoV-2 RT-qPCR, RNA was extracted from saline solution 0.9% with an automated method using magnetic beads (Sample Preparation System RNA, Abbott, Illinois, USA). SARS-CoV-2 RT-qPCR was performed using an adapted protocol described by Corman, Victor M et al, 2020 [10] to detect E gene as the first-line screening tool, followed by confirmatory testing with an assay detecting the N gene (Abbott, USA) and the commercial SARS-CoV-2 N1+N2 RT-qPCR kit to detect N1 and N2 genes (Qiagen, USA). SARS-CoV-2 RT-qPCR result was considered positive with an amplification cycle threshold (Ct) ≤ 32 and (Ct) ≤ 33, respectively.

Positive samples underwent genotyping for variant identification using TaqPathTM 1-Step RT-qPCR Master SARS-CoV-2 Mutation Panel Assay (Thermo Fisher Scientific, Waltham, USA). Data was analyzed by QuantStudio™ v2.5.1 design and analysis software in the genotyping module following IFU. The mutation panel was customized to detect the variants: Alpha (P681H), Beta (E484k + K417N), Gamma (E484K + T20N), Delta and Kappa (L425R + P681R), Zeta (E484K) and Lambda (L452Q).

### United Kingdom (UK)

In the UK, adults presenting with symptoms of COVID-19 (fever, cough, shortness of breath, tight chest, runny nose, sore throat, anosmia, ageusia, headache, diarrhoea, and tiredness) at a national community testing facility, the Liverpool John Lennon Airport drive-through COVID-19 test centre, were asked to participate in the study. Participants were recruited between July and August of 2021 under the Facilitating Accelerated COVID-19 Diagnostics (FALCON) study. Ethical approval was obtained from the National Research Ethics Service and the Health Research Authority (IRAS ID:28422, clinical trial ID: NCT04408170).

Swabs were taken systematically, NP swab samples in UTM (Copan Diagnostics Inc, Italy) were collected for the reference RT-qPCR test, this was followed by an NP swab to perform the Ag-RDTs. Due to biosafety restrictions at the drive-through centre, Ag-RDT testing were not done immediately after sample collection as per the IFU. All samples were transported in cooler boxes to the Liverpool School of Tropical Medicine (LSTM) and processed upon arrival by trained laboratory researchers following the IFU. Processing happened maximum within 3 hours of collection. Ag-RDTs were performed and the UTM NP swab samples were aliquoted and stored at -80°C until RNA extraction. RNA was extracted using the QIAamp^®^ 96 Virus QIAcube^®^ HT kit (Qiagen, Germany) on the QIAcube^®^ (Qiagen, Germany) and screened using TaqPath COVID-19 (ThermoFisher, UK) on the QuantStudio 5 TM thermocycler (ThermoFisher, UK). SARS-CoV-2 RT-qPCR result was considered positive if any two of the three targets (N, ORFab and S) were amplified with cycle threshold (Ct) ≤ 40.

### Analytical Sensitivity (UK only)

Viral culture methods to propagate SARS-CoV-2 isolates and to calculate plaque forming units per millilitre (PFU/mL) followed that previously described [11]. Briefly, isolates of SARS-CoV-2 from the wild type (Pango, B1) (REMRQ0001/Human/2020/Liverpool, GISAID ID EPI_ISL_464183), Alpha (B.1.1.7) (SARS-CoV-2/human/GBR/FASTER_272/2021, GenBank ID MW980115), Delta (B.1.617.2) (SARS-CoV-2/human/GBR/Liv_273/2021, GenBank ID OK392641), Gamma (P.1) (hCoV-19/Japan/TY7-503/2021, GISAID ID EPI_ISL_792683) and Omicron (B.1.1.529) (hCoV-19/USA/MD-HP20874/2021, GISAID ID EPI_ISL_7160424) lineages were used to evaluate the limit of detection (LOD) of the OnSite Ag-RDT. For the determination of the LOD, a fresh aliquot was serially diluted from 1.0× 10^5^ plaque forming units (pfu)/mL to 1.0 × 10^2^ pfu/mL. Each dilution was tested in triplicate. Two-fold dilutions were made below the ten-fold LOD dilution to confirm the lowest LOD (LLOD).

Viral RNA was extracted from each dilution using QIAmp Viral RNA mini kit (Qiagen, Germany) according to the manufacturer’s instructions, and quantified using Genesig RT-qPCR (Primer Design, UK). Genome copy number/mL (gcn/mL) were calculated as previously described [12].

### Statistical Analysis

The sensitivity and specificity, with 95% confidence intervals (CIs) were calculated based on the results of the reference method by RT-qPCR assay. Statistical analyses were performed using R scripts, Epi Info and GraphPad Prism 9.1.0 (GraphPad Software, Inc, California). The 95% confidence interval (CI) for the sensitivity and specificity was calculated using Wilson’s method. Fisher’s exact and chi-squared tests were used to determine non-random associations between categorical variables. Statistical significance was set at < 0.05.

## Results

### Clinical Evaluation

The demographics of both the Brazilian and UK study cohorts are shown in Table 1. In Brazil the median days from onset of symptoms was 3 [Q1-Q3, 2-4], with a vaccination rate of 97.0% (including partial and fully vaccinated participants). In the UK the median days from symptom onset was 2 [Q1-Q3, 1-3] and vaccination rate was 84.4% (including partial and fully vaccinated participants). Significantly higher SARS-CoV-2 RT-qPCR positivity was detected in the UK (36.5%, CI 0.29-0.43) than in Brazil (6.5%, CI 0.05-0.09) (*P <* 0.05).

**Table 1.** Demographics of Ag-RDT clinical evaluation cohorts for Brazil and United Kingdom.

| **Country** | **Brazil** | **United Kingdom** |
| --- | --- | --- |
| Age [mean (min-max), N] | 38.1 (16-69), 496 | 40.8 (20-86), 211 |
| Gender [%F, (n/N)] | 71.5 % (354/495) | 52.1% (110/210) |
| Symptoms present [%Yes, n/N] | 99.6% (494/495) | 100% (211/211) |
| Days from symptom onset [median (Q1-Q3); N] | 3 (2-4), 494 | 2 (1-3), 211 |
| Days 0-3 (n, %) | 294, 60% | 169, 80% |
| Days 4-7 (n, %) | 186, 38% | 36, 17% |
| Days 8+ (n, %) | 14, 3% | 6, 3% |
| Vaccinated (n, %) | 460, 93% | 132, 62% |
| Partially Vaccinated (n, %) | 19, 4% | 47, 22% |
| Not vaccinated (n, %) | 10, 2% | 32, 15% |
| Vaccination not disclosed (n, %) | 7, 1% | 1, 1%% |
| SARS-CoV-2 Positivity [%, (n/N)] | 6.5%, (32/496) | 36.5%, (77/211) |

The clinical sensitivity of the Onsite Ag-RDT across evaluation sites was heterogeneous, with the clinical sensitivity of 90.3% [95% Cl 75.1-96.7%] in Brazil and 75.3% [95% Cl 64.6-83.6%] in the UK (Table 2). The difference in sensitivities between sites was not statistically significant (*P* = 0.128). The clinical specificity of the Onsite Ag-RDT was 99.4% [95% Cl 98.1-99.8%] in Brazil and 95.5% [95% Cl 90.6-97.9%] in the UK.

**Table 2.** Results and clinical sensitivity and specificity of the OnSite COVID-19 Ag Device based on COVID-19 RT-qPCR result in Brazil and the United Kingdom.

| Results of<br>OnSiteCOVID-19 Ag<br>Device | Brazil |  |  | United Kingdom |  |  |
| --- | --- | --- | --- | --- | --- | --- |
|  | Confirmed by RT-qPCR |  |  |  |  |  |
|  | Positive | Negative | Total | Positive | Negative | Total |
| Positive | 28 | 3 | 31 | 58 | 6 | 64 |
| Negative | 3 | 461 | 464 | 19 | 128 | 147 |
| Total | 31 | 464 | 495 | 77 | 134 | 211 |
| Clinical Sensitivity<br>(95% CI), N | 90.3% (75.1-96.7%), 31 |  |  | 75.3% (64.6-83.6%), 77 |  |  |
| Clinical Specificity<br>(95% CI), N | 99.4% (98.1-99.8%), 464 |  |  | 95.5% (90.6-97.9%), 134 |  |  |
| Invalid Rate (% , n/N) | 0.6% (3/496) |  |  | 0% (211/211) |  |  |

RT-qPCR = Real-time quantitative polymerase chain reaction, Ct = cycle threshold, CI = confidence interval

In Brazil, of the 496 participants included, 32 were SARS-CoV-2 RT-qPCR positive (6.5%) (see Table 2). Twenty-eight of the RT-qPCR positive samples (90.3%) were Ag-RDT positive, while 3 (9.7%) were Ag-RDT negative and one was invalid (3.1%). Invalid results were removed for further analysis. Of the 464 RT-qPCR negative samples, 3 were Ag-RDT positive (0.6%). The sensitivity and specificity of the OnSite Ag-RDT test on RT-qPCR was 90.3% [95% Cl 75.1%-96.7%] and 99.4% [95% Cl 98.1%-99.8%], respectively (See Table 2). Sensitivity ≤7 days symptom onset was 96.2% [95% Cl 81.1-99.3%]. Sensitivity according to Ct value was 95% [95% Cl 75.1-99.8%] for Ct ≤25 and 90.3% [95% Cl 75.1%-96.7%] for Ct ≤33 (See Table 3). No statistical significance was found in sensitivity between different Ct value groups.

**Table 3.** COVID-19 RT-qPCR result in Brazil and the United Kingdom.

|  | **Brazil** | **United Kingdom** |
| --- | --- | --- |
| PCR Ct [median (Q1-Q3); N] | 19.6 (17.52-23), 31 | 19.5 (17.3-22.8), 77 |
| Ct > 33 (n, %) | 0, 0% | 1, 1% |
| Ct > 30 (n, %) | 1, 3% | 5, 6% |
| Ct > 25 (n, %) | 7, 22% | 11, 14% |
| Sensitivity Ct ≤20, N | 100%, (76.8-100%), 14 | 90.5% (77.4-97.3%), 42 |
| Sensitivity Ct ≤25, N | 95.0% (75.1-99.8%), 20 | 80.3% (69.2-88.1%), 66 |
| Sensitivity Ct ≤33, N | 90.3% (75.1-96.7%), 31 | 76.3% (65.5-84.5%), 76 |
| Sensitivity Ct ≤40, N | NA* | 75.3% (64.6-83.6%), 77 |

\* Maximum RT-qPCR cut off was ≤33 in Brazil

In the UK, of the 211 participants recruited, 77 (36.5%) were SARS-CoV-2 RT-qPCR positive (see Table 2). Fifty-Eight (75.3%) of the 77 RT-qPCR positive samples were also Ag-RDT positive, while 19 (24.7%) were Ag-RDT negative. Of the 134 RT-qPCR negative samples, 128 (95.5%) were also Ag-RDT negative and 6 (4.5%) were Ag-RDT positive. For the UK evaluation, the sensitivity and specificity were 75.3% [95% Cl 64.6-83.6%] and 95.5% [95% Cl 90.6-97.9%], respectively. Sensitivity ≤7 days symptom onset was 76.7% [95% Cl 65.8-84.9%]. Ct values of ≤20, ≤25, ≤33 and ≤40 had a sensitivity of 90.5% [95% Cl 77.4%-97.3%], 80.3% [95% Cl, 69.2%-88.1%], 76.3% [95% Cl 65.5-84,5%] and 75.3% [95% Cl 64.6-83.6%] respectively. Sensitivity was statistically higher among samples with Ct values ≤20 compared with samples with Ct values ≤33 (*P =* 0.029) and ≤40 (*P =* 0.044).

Subgroup analyses of the Brazilian and UK evaluation cohorts (Table 4) were performed to determine any associated differences in sensitivity compared to vaccination status and days from symptom onset. In the Brazilian cohort, sensitivity of the OnSite Ag-RDT was significantly lower on samples from patients with symptoms onset >7 days compared to samples with 0-3 symptoms onset (*P* = 0.02924) and samples with 0-7 days of onset (*P* = 0.03115) but no differences in sensitivity was found between groups of different vaccination status. In the UK, no difference in sensitivity was observed between groups of different symptoms onset and vaccination status (all *P* values >0.05). In Brazil, 52% of the positive samples were classified as Delta and 39% Gamma. In the UK, variant determination was not performed but at the time of enrollment, 100% of genome submissions corresponded to Delta variant [13].

**Table 4.** Ag-RDT result by onset of symptoms, and vaccinated individuals in Brazil and the UK.

|  | Brazil |  |  |  | United Kingdom |  |  |  |
| --- | --- | --- | --- | --- | --- | --- | --- | --- |
|  | Ag-RDT Positive (n, %) | Ag-RDT Negative (n, %) | Sensitivity<sup>a</sup> | 95% CI | Ag-RDT Positive (n, %) | Ag-RDT Negative (n, %) | Sensitivity<sup>a</sup> | 95% CI |
| **Days from symptom onset** |  |  |  |  |  |  |  |  |
| Days 0-3 | 16, 5.4% | 278, 94.6% | 100.0% | 76.9-100.0% | 52, 30.6% | 117, 69.4% | 79.7% | 67.2-89.0% |
| Days 4-7 | 12, 6.4% | 173, 93.6% | 91.7% | 61.5-99.8% | 10, 27.8% | 26, 72.2% | 64.3% | 35.2-87.3% |
| Days 8+ | 3, 21.4% | 11, 78.6% | 60.0% | 14.7-94.7% | 2, 33.3% | 4, 66.7% | 66.7% | 9.4-99.2% |
| **Vaccination received** |  |  |  |  |  |  |  |  |
| Vaccinated<sup>b</sup> | 31, 6.5% | 447, 93.5% | 93.3% | 77.4-99.2% | 52, 29.3% | 126, 70.7% | 78.3% | 65.8-87.9% |
| Not vaccinated | 1, 10.0% | 9, 90.0% | 0.00% | N/A | 11, 34.4% | 21, 65.6% | 62.5% | 35.4-84.8% |
| Not disclosed | 0, 0.0% | 7, 100.0% | N/A | N/A | 1, 100.0% | 0, 0.0% | 100% | 2.5-100.0% |

<sup>a</sup> As compared to RT-qPCR<sup>b</sup> Vaccinated defined as 1 or more doses
RT-qPCR = Real-time quantitative polymerase chain reaction, CI = confidence interval

### Analytical sensitivity

The LOD of the OnSite Ag-RDT was 1.0×10^3^ pfu/mL, 1.0×10^3^ pfu/mL, 1.0×10^2^ pfu/mL, 5.0×10^3^ pfu/mL and 1.0×10^3^ pfu/mL when tested on the WT, Alpha, Delta, Gamma and Omicron lineages, respectively. This gave a viral copy equivalent of approximately 2.1×10^5^ copies/mL, 2.1×10^4^ copies/mL, 1.6×10^4^ copies/mL 3.5×10 ^6^copies/mL and 8.7×10 ^4^copies/mL for the Ag-RDT for the WT, Alpha, Delta, Gamma and Omicron lineages.

## Discussion

The study aimed to evaluate the diagnostic performance of the OnSite COVID-19 Ag Rapid Test (CTK Biotech) in two different settings. Evaluating rapid diagnostic tests in diverse populations is vital to improving diagnostic responses as it gives an indication of the diagnostic accuracy in real world scenarios. In the case of rapid diagnostic testing within this pandemic, lateral flow tests which meet the minimum requirements for sensitivity and specificity can play a key role in increasing testing capacity, allowing timely clinical management of those infected and protecting healthcare systems [14]. This is particularly valuable in settings where access to the gold-standard RT-qPCR is often not available. Ag-RDTs are low cost, easy to use and do not require specialised skills or equipment which is essential to promote universal access.

Sensitivity and specificity of the OnSite Ag-RDT in a hospital setting in Brazil fulfilled the performance criteria determined by the World Health Organisation (WHO). However, sensitivity obtained in a community setting at a drive-through testing site in the UK missed the minimum recommendations [15] for both sensitivity and specificity. In guidance published by the WHO, minimum performance requirements for an Ag-RDT include a sensitivity of >80% and specificity of >97% [15]. Analytical evaluation of OnSite Ag-RDT detected Wild Type, Alpha, Delta and Omicron, meeting the recommendations in the WHO Target Product Profile for SARS-CoV-2 Ag-RDT of an acceptable analytical LOD of 1.0 × 10^6^RNA copies/mL [16] with the Gamma variant, slightly outside this threshold. In the Brazilian cohort, the Gamma variant was responsible for 39% of infections and the Delta variant was responsible for 52%. This is an interesting finding as it does not reflect the wider variant circulation in Brazil during this period as the Gamma variant was responsible for over 93% of infections in July 2021 and 70% of infections in August 2021 followed by Delta at 5% rising to 29% respectively [17]. In the UK, positive RT-qPCR results were not sequenced but it is assumed that all infections were Delta (B.1.617.2) due to the >99% circulation of this variant in the UK during the time of collection [18].

This study has several strengths, it is a multicentre and multinational evaluation across two different settings with differing testing capacity, prevalence of SARS-CoV-2 and population characteristics. In Brazil, samples were taken from a very exclusive population, healthcare workers in a healthcare setting with a high vaccination uptake compared to the rest of the population [19]. In the UK, data was collected from a diverse population, any person over the age of 18 presenting with COVID-19 symptoms at a government-run, drive-through COVID-19 testing facility. It is important to evaluate Ag-RDTs in a heterogeneous population and setting to obtain meaningful diagnostic accuracy data.

The main limitation for the study is that the drive-through testing setting in the UK did not allow for Ag-RDT testing to be performed at the point-of-care just after sample collection as recommended by the IFU. Guidance in the UK restricted testing of suspected COVID-19 positive individuals to high containment laboratories. Currently, there are limited studies on stability of Ag-RDT’s. In a systematic review on Ag-RDTs did not find a significant difference between Ninety-six data sets involved fresh specimens for antigen testing, and 23 data sets included freeze/thawed specimens for antigen testing [20] Although it is not stated whether the swabs were frozen dried or using transport buffer. However, one review of Ag-RDT performance in sub-Saharan Africa suggested that a delay in performing the test (CORIS COVID-19 Ag Respi-strip) may impact its stability if stored at 4^° C^ rather than frozen at -20^° C^ immediately [21]. Conversely, studies have shown that SARS-CoV-2 RNA remains stable for up to 9 days in dry swabs at ambient temperature of 20°C [22] and proteins are shown to be more stable than RNA [23]. Therefore, further investigation must take place to determine whether time from sample collection to Ag-RDT testing has a significant impact on the sensitivity.

Two other limitations of this study are that the RT-qPCR methodologies varied between both cohorts and the differences in SARS-CoV-2 prevalence. These factors have been attributed to a major cause of index case diagnostic accuracy [24]. For future evaluations, quantification of the viral copy numbers rather than Ct values is recommended to mitigate differences on RT-qPCR assay performances. This Ct variability has been estimated to be > 1000-fold in viral copy numbers/mL [24], as the RT-qPCR used in the UK has an LOD 10-fold more sensitive (10 genome copies/mL) than the RT-qPCR used in Brazil (100 genome copies/mL) [25]. The higher sensitivity of the RT-qPCR assay used in the UK, together with the higher cut-off used (Ct 40 versus Ct 32-33 in Brazil) could have contributed to higher numbers of false negatives in the index test compared to the Brazilian cohort. Additionally, there is a significant difference in sample size and in confirmed RT-qPCR positives (SARS-CoV-2 prevalence) between the two cohorts, with a low number of positive samples found in the Brazilian evaluation (6.3%) compared to the UK (36.5%). It has been reported that differences in prevalence can have an effect on the sensitivity and specificity of index tests [26, 27].

In conclusion, the data indicates that OnSite Ag-RDT had lower performance quality than published by the manufacturers for the detection of SARS-CoV-2 in clinical samples and varied greatly between the two settings in this study. Further evaluation of the use of Ag-RDTs should strictly follow the IFUs of the test and include harmonised protocols between laboratories to facilitate comparison between settings. In particular, the use of viral copy numbers rather than Ct values has been suggested to minimise the variability between laboratories.

## Data Availability

All data produced in the present study are available upon reasonable request to the authors

## Acknowledgments

LSTM diagnostics group: Ms Caitlin Greenland Bews, Ms Kate Buist, Ms Karina Clerkin, Dr Thomas Edwards, Dr Lorna Finch, Dr Helen Savage, Ms Jahanara Wardale, Ms Rachel Watkins, Mr Chris Williams and Mr Dominic Wooding.

Condor steering group: Dr A. Joy Allen, Dr Julian Braybrook, Professor Peter Buckle, Ms Eloise Cook, Professor Paul Dark, Dr Kerrie Davis, Dr Gail Hayward, Professor Adam Gordon, Ms Anna Halstead, Dr Charlotte Harden, Dr Colette Inkson, Ms Naoko Jones, Dr William Jones, Professor Dan Lasserson, Dr Joseph Lee, Dr Clare Lendrem, Dr Andrew Lewington, Mx Mary Logan, Dr Massimo Micocci, Dr Brian Nicholson, Professor Rafael Perera-Salazar, Mr Graham Prestwich, Dr D. Ashley Price, Dr Charles Reynard, Dr Beverley Riley, Professor John Simpson, Dr Valerie Tate, Dr Philip Turner, Professor Mark Wilcox, Dr Melody Zhifang.

We would like to acknowledge the participants for volunteering for this study and to the CRN for supporting us with the sample collection and recruitment during the study; particularly to Sue Dowling and Larysa Mashenko; we also acknowledge the support of the UK National Institute for Health Research Clinical Research Network and the COvid-19 National DiagnOstic Research & evaluation (CONDOR) programme. In Brazil, we would like to thank the participants and the Centro de atendimento ao colaborador, Hospital das Clínicas da Faculdade de Medicina da Universidade de São Paulo, São Paulo, Brazil.

## Research Funding

This work was funded as part of FIND’s work as co-convener of the diagnostics pillar of the Access to COVID-19 Tools (ACT) Accelerator, including support from Unitaid [grant number: 2019-32-FIND MDR], the governments of the Netherlands [grant number: MINBUZA-2020.961444] and from UK Department for International Development [grant number 300341-102]. The FALCON study was funded by the National Institute for Health Research, Asthma UK and the British Lung Foundation. This work is partially funded by the National Institute for Health Research (NIHR) Health Protection Research Unit in Emerging and Zoonotic Infections (200907), a partnership between the UK Health Security Agency, The University of Liverpool, The University of Oxford and The Liverpool School of Tropical Medicine. The views expressed are those of the author(s) and not necessarily those of the NIHR, the UKHSA or the Department of Health and Social Care.

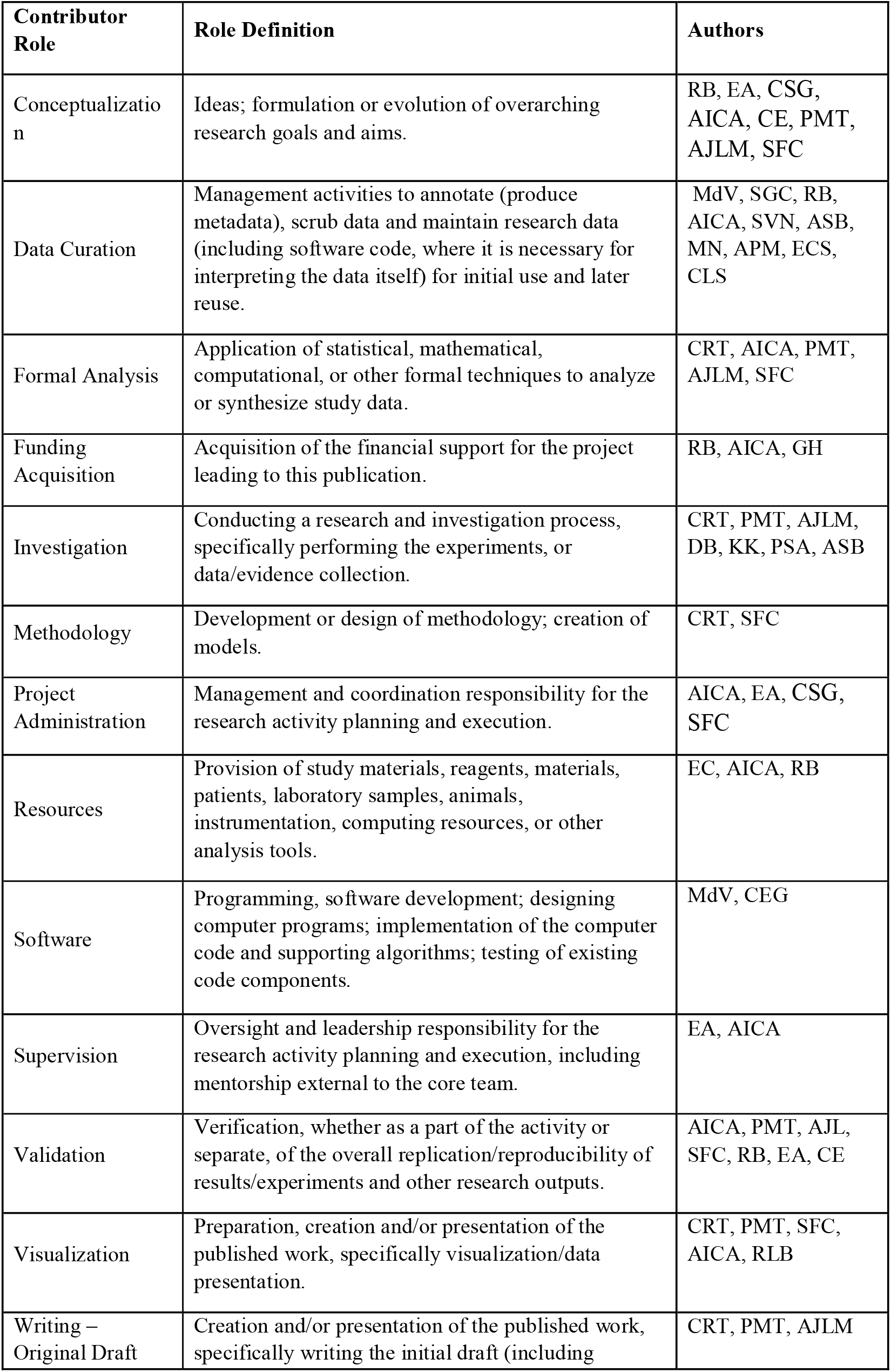
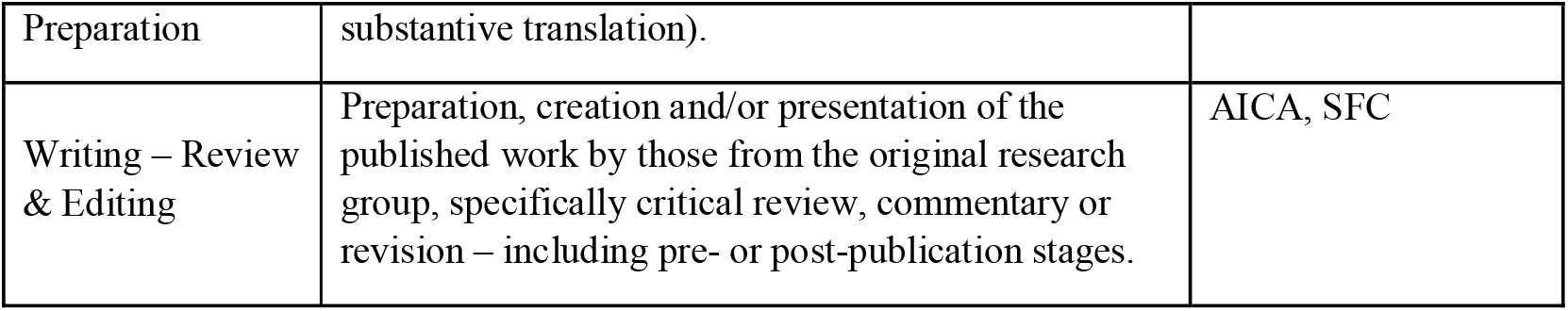

## References

1. FIND. Test Directory,. 2022; Available from: https://www.finddx.org/test-directory/?_test_format=lateral-flow-assay-strip-or-cassette&_assay_target=antigen.

2. Dinnes, J., et al., Rapid, point-of-care antigen and molecular-based tests for diagnosis of SARS-CoV-2 infection. Cochrane Database of Systematic Reviews, 2021(3).

3. Krüger, L.J., et al., Accuracy and ease-of-use of seven point-of-care SARS-CoV-2 antigen-detecting tests: A multi-centre clinical evaluation. eBioMedicine, 2022. 75: p. 103774.

4. World Health Organization, Recommendations for national SARS-CoV-2 testing strategies and diagnostic capacities. 2021.

5. Kameda, K., et al., Testing COVID-19 in Brazil: fragmented efforts and challenges to expand diagnostic capacity at the Brazilian Unified National Health System. Cad Saude Publica, 2021. 37(3): p. e00277420.

6. Pilkington, V., S.M. Keestra, and A. Hill, Global COVID-19 Vaccine Inequity: Failures in the First Year of Distribution and Potential Solutions for the Future. Frontiers in Public Health, 2022. 10.

7. Mathieu, E., et al., A global database of COVID-19 vaccinations. Nature Human Behaviour, 2021. 5(7): p. 947–953.

8. World Health Organization. Tracking SARS-CoV-2 variants. 2022 [cited 2022 28/01/2022]; Available from: https://www.who.int/en/activities/tracking-SARS-CoV-2-variants/.

9. Berti, L. Patients in Sao Paulo waiting a week for Covid-19 test results. 2022; Available from: https://brazilian.report/liveblog/2022/01/21/patients-sao-paulo-test-results/.

10. Corman, V.M., et al., Detection of 2019 novel coronavirus (2019-nCoV) by real-time RT-PCR. Eurosurveillance, 2020. 25(3): p. 2000045.

11. Edwards, T., et al., SARS-CoV-2 Transmission Risk from sports Equipment (STRIKE). medRxiv, 2021: p. 2021.02.04.21251127.

12. Cubas-Atienzar, A.I., et al., Limit of detection in different matrices of 19 commercially available rapid antigen tests for the detection of SARS-CoV-2. Scientific Reports, 2021. 11(1): p. 18313.

13. Next Strain. Genomic epidemiology of novel coronavirus - Global subsampling. 2022; Available from: https://nextstrain.org/ncov/gisaid/global?f_country=United%20Kingdom.

14. Peeling, R.W., et al., Diagnostics for COVID-19: moving from pandemic response to control. The Lancet, 2022. 399(10326): p. 757–768.

15. World Health Organziation, Coronavirus disease 2019 (COVID-19): situation report,. 2020, World Health Organization.

16. World Health Organization, COVID-19 Target product profiles for priority diagnostics to support response to the COVID-19 pandemic v.1.0. 2020.

17. Our World Data. SARS-CoV-2 variants in analyzed sequences. 2022; Available from: https://ourworldindata.org/grapher/covid-variants-area?country=~BRA.

18. Our World Data. SARS-CoV-2 variants in analyzed sequences, United Kingdom. 2022 [cited 2022; Available from: https://ourworldindata.org/grapher/covid-variants-area?country=~GBR.

19. Coronavirus Brazil. Painel Coronavírus Atualizado. 2022; Available from: https://covid.saude.gov.br/.

20. Parvu, V., et al., Factors that Influence the Reported Sensitivity of Rapid Antigen Testing for SARS-CoV-2. Frontiers in Microbiology, 2021. 12.

21. Jacobs, J., et al., Implementing COVID-19 (SARS-CoV-2) Rapid Diagnostic Tests in Sub-Saharan Africa: A Review. Front Med (Lausanne), 2020. 7: p. 557797.

22. Gokulan, C.G., et al., Temporal stability and detection sensitivity of the dry swab-based diagnosis of SARS-CoV-2. Journal of Biosciences, 2021. 46(4): p. 95.

23. How fast do RNAs and Proteins degrade? Cell biology by the numbers; Available from: http://book.bionumbers.org/how-fast-do-rnas-and-proteins-degrade/.

24. Evans, D., et al., The Dangers of Using Cq to Quantify Nucleic Acid in Biological Samples: A Lesson From COVID-19. Clin Chem, 2021. 68(1): p. 153–162.

25. Sohni, Y., Variation in LOD Across SARS-CoV-2 Assay Systems: Need for Standardization. Laboratory Medicine, 2020. 52(2): p. 107–115.

26. Leeflang, M.M.G., P.M.M. Bossuyt, and L. Irwig, Diagnostic test accuracy may vary with prevalence: implications for evidence-based diagnosis. Journal of Clinical Epidemiology, 2009. 62(1): p. 5–12.

27. Leeflang, M.M., et al., Variation of a test’s sensitivity and specificity with disease prevalence. Cmaj, 2013. 185(11): p. E537–44.

